# Disentangling mechanisms behind the pleiotropic effects of proximal 16p11.2 BP4-5 CNVs

**DOI:** 10.1101/2024.03.20.24304613

**Authors:** Chiara Auwerx, Samuel Moix, Zoltán Kutalik, Alexandre Reymond

## Abstract

Whereas 16p11.2 BP4-5 copy-number variants (CNVs) represent one of the most pleiotropic etiologies of genomic syndromes in both clinical and population cohorts, the mechanisms leading to such pleiotropy remain understudied. Identifying 73 deletion and 89 duplication carriers among unrelated white British UK Biobank participants, we performed a phenome-wide association study between the region’s copy number and 117 complex traits and diseases, mimicking four dosage models. Forty-six phenotypes (39%) were affected by 16p11.2 BP4-5 CNVs, with the deletion-only, mirror, U-shape, and duplication-only models being the best fit for thirty, ten, four, and two phenotypes, respectively, aligning with the stronger deleteriousness of the deletion. Upon individually adjusting CNV effects for either body mass index (BMI), height, cognitive function, or socio-economic status as potential mediators, we found that sixteen testable deletion-driven associations (61%) – primarily with cardiovascular and metabolic traits – were BMI-dependent, with other mediators playing a more subtle role. Bidirectional Mendelian randomization supported that 13 out of these 16 associations (81%) were secondary consequences of the CNV’s impact on BMI. For the 22 traits that remained significantly associated upon individual adjustment for mediators, matched-control analyses found that eleven phenotypes, including musculoskeletal traits, liver enzymes, fluid intelligence, platelet count, pulmonary capacity, pneumonia, and acute kidney injury, remained associated under strict Bonferroni correction, with eight additional nominally significant associations. These results paint a complex picture of 16p11.2 BP4-5’s pleiotropic pattern that involves direct effects on multiple physiological systems and indirect co-morbidities consequential to the CNV’s impact on BMI and cognition, acting through trait-specific dosage mechanisms.

## Introduction

Genomic disorders are caused by recurrent genomic rearrangements that lead to the gain (duplication) or loss (deletion) of large, multi-kilobase pair (kb) DNA fragments. The proximal 16p11.2 rearrangement spans a region of ∼600 kb between recurrent breakpoints (BP) 4 and 5 and includes 27 unique protein-coding genes. Copy-number variants (CNVs) of the region represent one of the most common genomic disorders, with population prevalence estimates of 1 in 3,000 and 1 in 2,800 for the deletion (MIM: 611913) and duplication (MIM: 614671), respectively (*in litt.*). Prevalence in clinical cohorts is about eight-fold higher, with a particularly strong enrichment in individuals ascertained for intellectual disability and developmental delay^1–3^ or autism spectrum disorder^4–7^, the first phenotypes associated with the CNV. Other hallmark features include a negative dosage effect on body mass index (BMI)^8–10^ and head circumference^11,12^, a predisposition for seizure disorders^2,3,11,13^, and a duplication-specific increased susceptibility to schizophrenia and other psychiatric conditions^12,14–19^. The recent establishment of large biobanks coupling genetic information to phenotypic data such as physical measurements, blood biomarkers, and electronic health records, has allowed to study the phenotypic expression of 16p11.2 BP4-5 rearrangements in individuals that are typically older and less severely affected than those recruited in pediatric clinical cohorts^20–31^. Results of these studies often converge onto similar pathophysiological processes than those highlighted by clinical studies but also report associations with biomarkers and common diseases that are typically overlooked or not assessed in clinical cohorts.

If the pleiotropic nature of 16p11.2 BP4-5 rearrangements is now well-established, the mechanisms through which CNVs in the region affect such diversity of traits remain poorly studied. Under a model of direct (or horizontal) pleiotropy, the CNV causally impacts associated phenotypes through independent mechanisms (Figure 1A). Conversely, indirect (or vertical) pleiotropy implies that the CNV causally impacts a mediatory trait, which in turn causally impacts other traits that will appear as linked with the CNV in association studies (Figure 1B). These models are not mutually exclusive, and a fraction of the associations might result from direct effects while others might be secondary consequences. This question is particularly relevant given the BMI-modulating role of the 16p11.2 BP4-5 CNV^8–10,20,21,24,25^ – which itself represents a strong risk factor for other diseases – and could therefore inform epidemiology of associated comorbidities and clinical practice. To address this knowledge gap, we re-analyzed two recent UK Biobank (UKBB) studies that assessed the impact of 16p11.2 BP4-5 rearrangements on 117 complex traits and common diseases^21,22^ with the aims to i) determine the most likely dosage mechanism for different traits and ii) estimate the fraction and nature of associations that are mediated by primary changes in anthropometric measurements, cognitive ability, and socio-economic status (Figure 1C).

**Figure 1.**
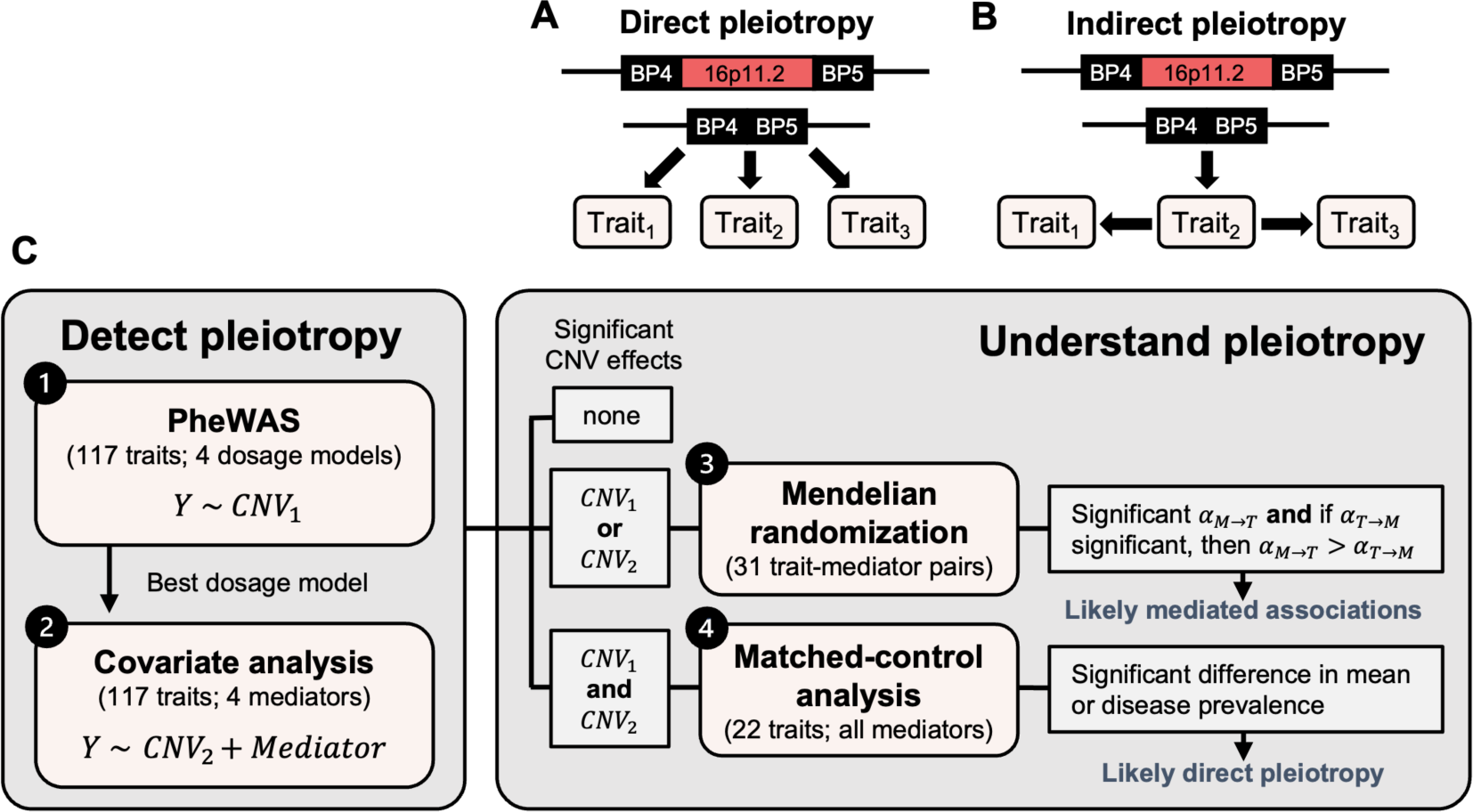
Study workflow. (**A**-**B**) Schematic representation of pleiotropy mechanisms. For illustration, the 16p11.2 BP4-5 deletion is depicted but the same concept applies to the duplication. (A) Direct (horizontal) pleiotropy: The CNV causally affects multiple traits – here Trait_1_, Trait_2_, and Trait_3_ – through independent mechanisms. (B) Indirect (vertical) pleiotropy: The CNV causally impacts Trait_2_, which in turn causally affects Trait_1_ and Trait_3_. The impact of the deletion on Trait_1_ and Trait_3_ is thus indirect and mediated by a shared mechanism, i.e., Trait_2_. (**C**) Overview of the study. The first two analyses aim at detecting and characterizing the pleiotropy of 16p11.2 BP4-5 CNVs through four distinct dosage models that estimate the effect of the CNV on the trait (Y) either (1) without or (2) with adjustment for one of four covariates that could potentially mediate the CNV-phenotype association. The second part of the study aims at understanding the mechanisms through which pleiotropy arises. (3) Bidirectional Mendelian randomization was used to investigate the causal relationship between trait-mediator pairs for which the significance of the CNV effect on the trait was affected by adjustment for the mediator. Support for mediation was claimed when the forward MR effect of the mediator on the trait (*α*_*M*→*T*_) is significant and larger than the reverse effect of the trait on the mediator (*α*_*T*→*M*_), providing the latter is significant. (4) For traits that showed a significant association with the CNV regardless of covariate adjustment, we performed a matched-control analysis that allowed us to adjust for all possible mediators simultaneously and detect likely genuine, direct pleiotropic associations. PheWAS = phenome-wide association study.

## Materials and methods

### 1. Study material

#### Software versions

Statistical analyses and graphs were generated with R v4.3.1. The Mendelian randomization analysis pipeline uses the `TwoSampleMR` R package v0.5.7^32^ and PLINK v1.9^33^ and was implemented in R v4.2.1.

#### Cohort description & sample selection

Analyses were carried out in the UKBB, a volunteer-based UK population cohort of about half a million individuals (54% females) aged 40-69 years at recruitment, who signed a broad informed consent form^34^. Available data include microarray genotype data acquired in GRCh37/hg19 from two similar arrays, as well as rich phenotypic data, including anthropometric measurements, vital signs, blood biomarker levels, life history and lifestyle questionnaire data, hospital-based International Classification of Diseases, 10^th^ Revision (ICD-10) codes (up to September 2021), and self-reported conditions. Analyses conducted in this study focus on 331,522 unrelated individuals from the “white British” UKBB subset (54% females) that were filtered to exclude samples with abnormal CNV profiles and/or a report of blood malignancy. Filtering criteria to obtain this set are described elsewhere^22^.

#### CNV carrier identification

CNV calls from a previous study were used^21^. Briefly, CNV calling was done based on the UKBB microarray data using standard PennCNV v1.0.5 settings^35^. Each call was attributed a quality score ranging from −1 (likely deletion) to 1 (likely duplication) reflecting the probability for the CNV to be a consensus call across three algorithms and thus a true positive^36^. 16p11.2 BP4-5 deletion and duplication carriers were identified as carrying a high-confidence CNV call (quality score < −0.5 for deletions; quality score > 0.5 for duplications) on chromosome 16 with start and end site within 29.4-29.8 Mb and 30.05-30.4 Mb, respectively. Individuals with a low-quality 16p11.2 BP4-5 CNV were excluded from copy-neutral controls. CNV genotype vectors were then encoded to allow the fitting of regression models according to four dosage mechanisms (Table 1).

**Table 1.** Encoding of CNV carrier status for different dosage models. Numerical encoding of CNV genotypes for high confidence deletion carriers, copy-neutral individuals, and high confidence duplication carriers according to four dosage mechanisms of action. Individuals with a low-quality score CNV call are set as missing.

|  | deletion<br>(copy-number = 1) | copy-neutral<br>(copy-number = 2) | duplication<br>(copy-number = 3) |
| --- | --- | --- | --- |
| mirror | -1 | 0 | 1 |
| U-shape | 1 | 0 | 1 |
| duplication-only | NA | 0 | 1 |
| deletion-only | 1 | 0 | NA |

#### Phenotype selection

We analyzed the same 117 phenotypes as defined in previous studies^21,22^. This includes 57 quantitative traits^21^ that were inverse normal transformed before being corrected for sex (except for sex-specific traits), age (UKBB field identifier #21003), age^2^, genotyping batch, and principal components 1-40. We further include 60 common diseases based on ICD-10 clinical diagnoses using a case-control definition procedure that excludes from controls individuals with a condition related to the one under investigation^22^.

#### Mediator selection

We tested the role of four factors that could potentially mediate associations between 16p11.2 BP4-5 CNVs and the assessed phenotypes:

- Body mass index (BMI): average over available instances of BMI (#21001).
- Educational attainment (EA): age at which full-time education was completed (#845). Values matching “prefer not to answer”, “never went to school”, and “do not know” were set as missing, and average over available instances was calculated. Individuals for which average age at which full-time education was completed was below 14 years or over 19 years were set to 14 years and 19 years, respectively. Individuals reporting a “college or university degree” in their qualifications (#6138) were set to 19 years.
- Townsend deprivation index at recruitment (TDI; #22189).
- Height: average over available instances of standing height (#50).

#### GWAS summary statistics

Mendelian randomization (MR) studies rely on publicly available genome-wide association studies (GWASs) summary statistics for both sexes and individuals of European ancestry. For mediators, summary statistics from Pan-UK Biobank with phenotype manifest updated on 01/03/2023 (https://pan.ukbb.broadinstitute.org/)^37^ were used for BMI, TDI, and height. For EA, summary statistics from a large meta-analysis were used (excluding 23andMe data; http://www.thessgac.org/data)^38^. For other phenotypes, summary statistics from the Neale group released on 07/2018 were used (http://www.nealelab.is/uk-biobank). These summary statistics were favored over those of large disease-specific consortia as summary statistics for binary traits were calculated through linear regression, allowing comparison of forward and reverse effects. For diseases, we used the closest possible match to our phenotype definition, i.e., phenotype code: E10 for “T1D” (type 1 diabetes); G47 for “sleep” (sleep apnea); I10 for “HTN_essential” (essential hypertension); I35 for “valves” (cardiac valve disorders); I44 for “conduction” (cardiac conduction disorders); J45 for “asthma”; M19 for “OA” (arthrosis); N18 for “CKD” (chronic kidney disease); 20002_1473 for “lipid” (lipidemias & lipoprotein disorders). Summary statistics for autosomal chromosomes were harmonized to the UK10K reference panel^39^. After excluding palindromic single-nucleotide polymorphisms (SNPs) and adjusting strand-flipped SNPs, effect sizes were standardized to represent the square root of the explained variance.

### 2. 16p11.2 BP4-5 association studies

#### Phenome-wide association study

For the phenome-wide association study (PheWAS), regression analysis was performed to estimate the effect of the CNV genotype – encoded according to either of the four models in Table 1 – and the 117 selected phenotypes. For quantitative traits, linear regressions (`lm()` in R) were used and 95% confidence intervals (CI) were calculated as beta ± 1.96*standard error (SE). For binary traits, Firth’s bias-reduced penalized-likelihood logistic regression was used (`logistf(plconf = 2, maxit = 100, maxstep = 10)` from the `logistf` package v1.26.0 in R) to account for the fact that both CNV carriers and disease cases are rare. The same function also produces estimates for the 95% CIs. As disease diagnoses were defined as binary variables and could not be adjusted beforehand, sex (except for sex-specific traits), age, genotyping array, and principal components 1-40 were included as covariates. For each trait, the dosage model yielding the lowest p-value for the CNV effect was retained and effects were defined as strictly significant under Bonferroni correction criteria (p ≤ 0.05/117 = 4.3 x 10^−4^).

#### Covariate analysis

For all phenotype-mediator pairs (*Phenotype* & *Mediator selection*), including those involving phenotypes that did not significantly associate with the CNV status in our original PheWAS, we estimated the Pearson correlation (`cor(use = “pairwise.complete.obs”)` in R), as well as the effect of the mediator on the phenotype in a linear/Firth regression model without covariates, as previously described (*Phenome-wide association study*). For pairs with Pearson correlation < 0.5 and effect of the mediator on the trait p ≤ 0.05/117 = 4.3 x 10^−4^, we estimated the effect of the CNV carrier status encoded according to the best PheWAS model. Regressions were implemented as previously described (*Phenome-wide association study*), adding the mediator as an additional covariate. Adjusted effects were defined as strictly significant when meeting Bonferroni correction criteria (p ≤ 0.05/117 = 4.3 x 10^−4^). We additionally compared effect estimates with (*β_adjusted_*) and without (*β*) mediator adjustment based on a t-statistic

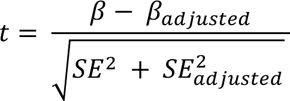

where SEs represent the standard errors of the effects. Two-sided p-values were calculated (`2*pnorm(-abs(t), mean = 0, sd = 1)` in R). The difference in correlation between BMI-dependent and BMI-independent traits with BMI was assessed with a two-sided t-test.

#### Mendelian randomization

GWAS summary statistics were used to conduct bidirectional MR according to a previously published pipeline^40,41^ for 31 mediator-trait pairs for which the CNV-trait association either gained or lost significance upon adjusting for that mediator. Concretely, the forward effect of the mediator (exposure) on the trait (outcome) and the reverse effect of the trait (exposure) on the mediator (outcome) were estimated. Harmonized SNPs significantly (p < 5 x 10^−8^) associating with the exposure were clumped with PLINK v1.9 (`p1 = 0.0001, p2 = 0.01, kb = 250, and r2 = 0.01`) and retained as instrumental variables. Instrumental variables mapping to the extended HLA region (chr6:25,000,000–37,000,000; GRCh37/hg19) were excluded, as well as those with a difference in allele frequency (≥ 0.05) between the outcome and exposure summary statistic. Steiger filtering was applied (Z ≤ −1.96) to ensure that the effect of the selected variants on the exposure was stronger than their effect on the outcome. Bidirectional inverse variance weighted MR analyses were carried out with the `TwoSampleMR` R package when at least two instrumental variables were available. MR effects were called significant under Bonferroni correction, when p ≤ 0.05/62 = 8.1 x 10^−4^, to account for the 31 bidirectional tests performed.

#### Matched-control analysis

For each CNV carrier, we identified all copy-neutral unrelated individuals from the “white British” subset of UKBB participants that were matching based on sex (identical), age (± 2.5 years), BMI (± 2.5 years), TDI (± 2), average household income before tax (#738) averaged over instances (identical category), and EA (± 1 year). Fifty-eight deletion and sixty-one duplication carriers had no missing data and qualified for the matching procedure. The number of identified matching controls per carrier ranged from 1 to 918 and 12 to 1,590 for deletion and duplication carriers, respectively, with 49 deletion and 60 duplication carriers having at least 25 matching controls. When more than 25 matched controls were available, the ones used for the analysis were selected randomly (`sample_n()` in R), without replacement. For quantitative traits, we compared mean phenotypic values between deletion and duplication carriers and the respective control groups through a two-sided t-test. For binary traits, disease prevalence was compared between the same groups based on a two-sided Fisher test. Prevalence standard error was calculated as

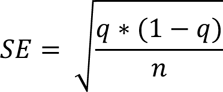

where *q* is the disease prevalence, and *n* the sample size. Sample sizes vary between phenotypes due to missing data. We define significant associations based on a Bonferroni correction that accounts for the 22 traits of interest in this analysis (p ≤ 0.05/22 = 2.3 x 10^−3^), i.e., phenotypes that remained associated with the CNV under strict Bonferroni correction when adjusting for BMI, height, EA, or TDI individually. We report all nominally significant (p < 0.05) associations on figures.

In a related analysis aiming at assessing the consequences of losing samples for the matched-control analysis, we used the same statistical framework to compare mean phenotypic value and disease prevalence between deletion and duplication carriers that were included in the matched-control analysis versus those that were not due to missing data or lack of sufficient controls.

## Results

Using previously published high confidence CNV calls for 331,522 unrelated, white British UKBB participants^21,22^, we identified 73 and 89 individuals with a 16p11.2 BP4-5 (start: chr16:29.40-29.80 Mb; end: chr16:30.05-30.40 Mb) deletion and duplication, respectively. CNV genotypes were encoded to allow testing of four dosage mechanisms, namely a mirror model assessing the additive impact of each additional copy, a U-shape model testing the same-direction impact of any deviation from the copy-neutral state, and duplication- and deletion-only models that assess the separate impact of duplications and deletions, respectively. Next, we evaluated the association between an individual’s CNV carrier status and 117 phenotypes – that comprise 57 quantitative variables including anthropometric measurements, vital signs, biomarker levels, life history events, and 60 common diseases – while correcting for sex, age, age^2^, genotyping array, and population stratification (Figure 2; Table S1).

**Figure 2.**
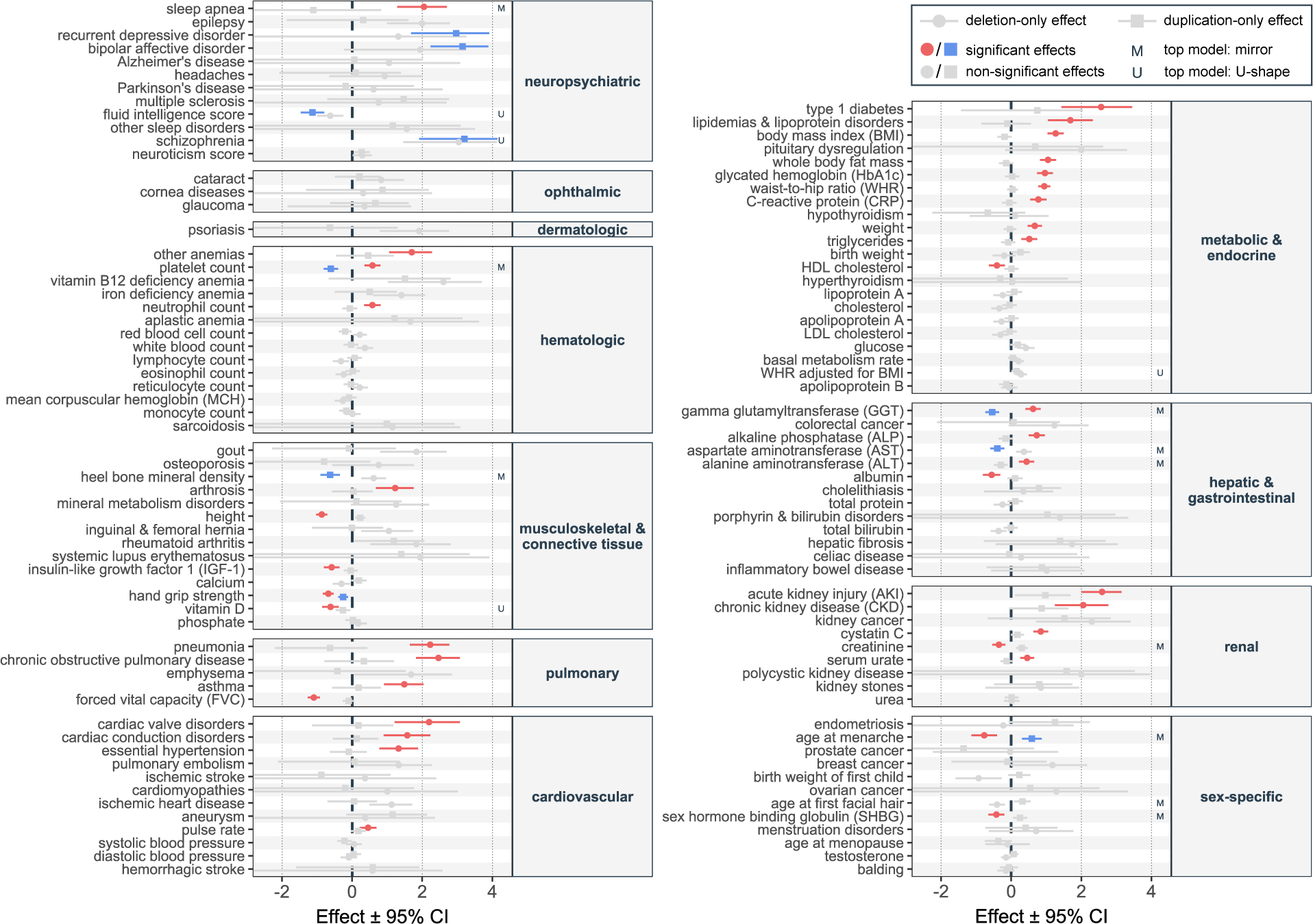
16p11.2 BP4-5 phenome-wide association study. Effect sizes (beta; x-axis) with 95% confidence interval (CI) of the 16p11.2 BP4-5 deletion (circle) and duplication (square) on 117 complex traits and diseases, ordered by physiological system (y-axis). Effect sizes are in standard deviation units of the outcome (quantitative traits) or logarithms of the odds ratio of a logistic regression (disease traits). Deletion- and duplication-only effects that are significant under Bonferroni correction (p ≤ 0.05/117 = 4.3 x 10^−4^) are in blue and red, respectively, while non-significant effects are in grey. If the most significant among the four tested association models was the mirror or U-shape model, it is denoted with an “M” or “U”, respectively (right).

Overall, 46 (39%) traits, including 16 diseases, were associated with the CNV carrier status under at least one association model (Bonferroni correction: p ≤ 0.05/117 = 4.3 x 10^−4^; Table 2), with an additional 32 (27%) showing a trend for association (nominal significance: p ≤ 0.05). Specifically, 10 and 38 traits showed a significant association through the duplication-only and deletion-only models, respectively, indicating a stronger propensity for pleiotropy and deleteriousness of the deletion, compared to the duplication. Exceptions are recurrent depression and bipolar disorder, the two only traits for which the duplication-only model yielded the most significant result. This is in line with the duplication representing a strong susceptibility factor for psychiatric conditions^12,14–19^. Similarly, the risk for schizophrenia was strongly increased by the duplication, even if our analysis finds that the relation is better described by a U-shape model wherein the deletion also tends to increase schizophrenia risk. Surprisingly, the CNV did not associate with neuroticism score, despite the high genetic correlation between neuroticism and psychiatric conditions^42^. Three other traits, namely fluid intelligence, vitamin D, and waist-to-hip ratio adjusted for BMI (WHRadjBMI), were also most significantly associated through a U-shape effect, while grip strength was decreased in both deletion and duplication carriers, but more strongly in the former. Conversely, ten traits were most significantly associated through a mirror model, including multiple hepatic biomarkers, platelet count, and traits related to sexual characteristics such as puberty timing and sex hormone binding globulin (SHBG) levels. Finally, the deletion-only model was the most significant fit for 30 phenotypes, including mostly pulmonary, cardiovascular, metabolic, and renal traits.

**Table 2.**
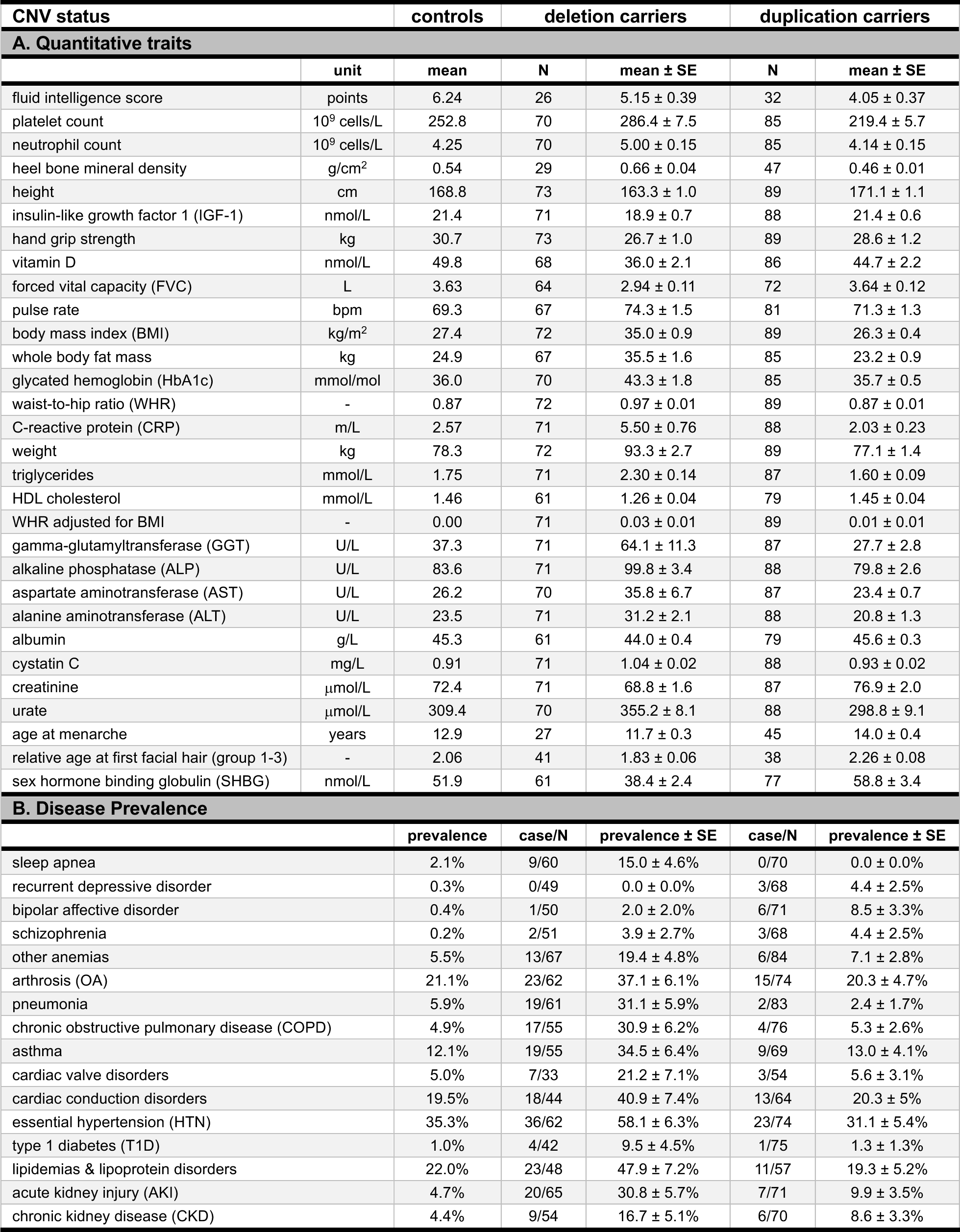
Traits significantly associated with 16p11.2 BP4-5 CNVs. Traits that are significantly (p ≤ 0.05/117 = 4.3 x 10^−4^) associated with 16p11.2 BP4-5 CNVs through at least one of the four assessed association models, following the ordering of Figure 2. (**A**) For quantitative traits, the mean value of the traits in copy-neutral individuals (controls) is provided along with the mean value and standard error (SE) among duplication and deletion carriers. The number of duplication and deletion carriers with available data is specified as N. Values are given in the indicated unit. (**B**) For binary disease traits, prevalence in percentage among copy-neutral individuals is provided along with prevalence and SE among duplication and deletion carriers. Diseased (case) and total (N) number of duplication and deletion carriers are indicated.

Having characterized the pleiotropic nature of 16p11.2 BP4-5 rearrangements, we next sought to establish whether some of these associations might be secondary to the CNV affecting core mediatory phenotypes, i.e., reflect indirect pleiotropy (Figure 1B). We focus on four traits that proxy hallmark features of the 16p11.2 BP4-5 rearrangement and have the potential to influence other associated traits: i) BMI, which characterizes the negative correlation between dosage and adiposity^8–10,20,21,24,25^ and represents a major risk factor for many common diseases; ii) Height, which is reduced in deletion carriers^16,20,21,24,36^ and can influence musculoskeletal phenotypes; iii) Educational attainment (EA) proxied by age at which an individual completed their education. This variable offers the advantage of being available for the near totality of the UKBB cohort while strongly correlating with fluid intelligence score that is limited to about half of its participants (Pearson correlation = 0.42), thereby reflecting the decreased cognitive function observed in both duplication and deletion carriers^2,3,16,17,21,31,43^; and iv) Townsend deprivation index (TDI) as a measure of socio-economic status (SES), which we expect to be reduced as a corollary of the health burden imposed by the CNV^20^. Of note, while TDI specifically aims at assessing SES, BMI, height, and EA also partly capture SES^44^. For the association between CNV and phenotype to be mediated by one of these factors, the mediator needs to significantly (p ≤ 0.05/117 = 4.3 x 10^−4^) associate with the tested phenotype. Furthermore, phenotypes cannot be too correlated with the mediator (Pearson’s correlation > 0.5), as in such situations distinguishing mediator and outcome would be particularly difficult. For all mediator-trait pairs that fulfilled these criteria, we tested the impact of adjusting the CNV-trait effect for mediatory factors by including them individually in the regression model yielding the most significant CNV-trait effect (Figure 3A; Table S2).

**Figure 3.**
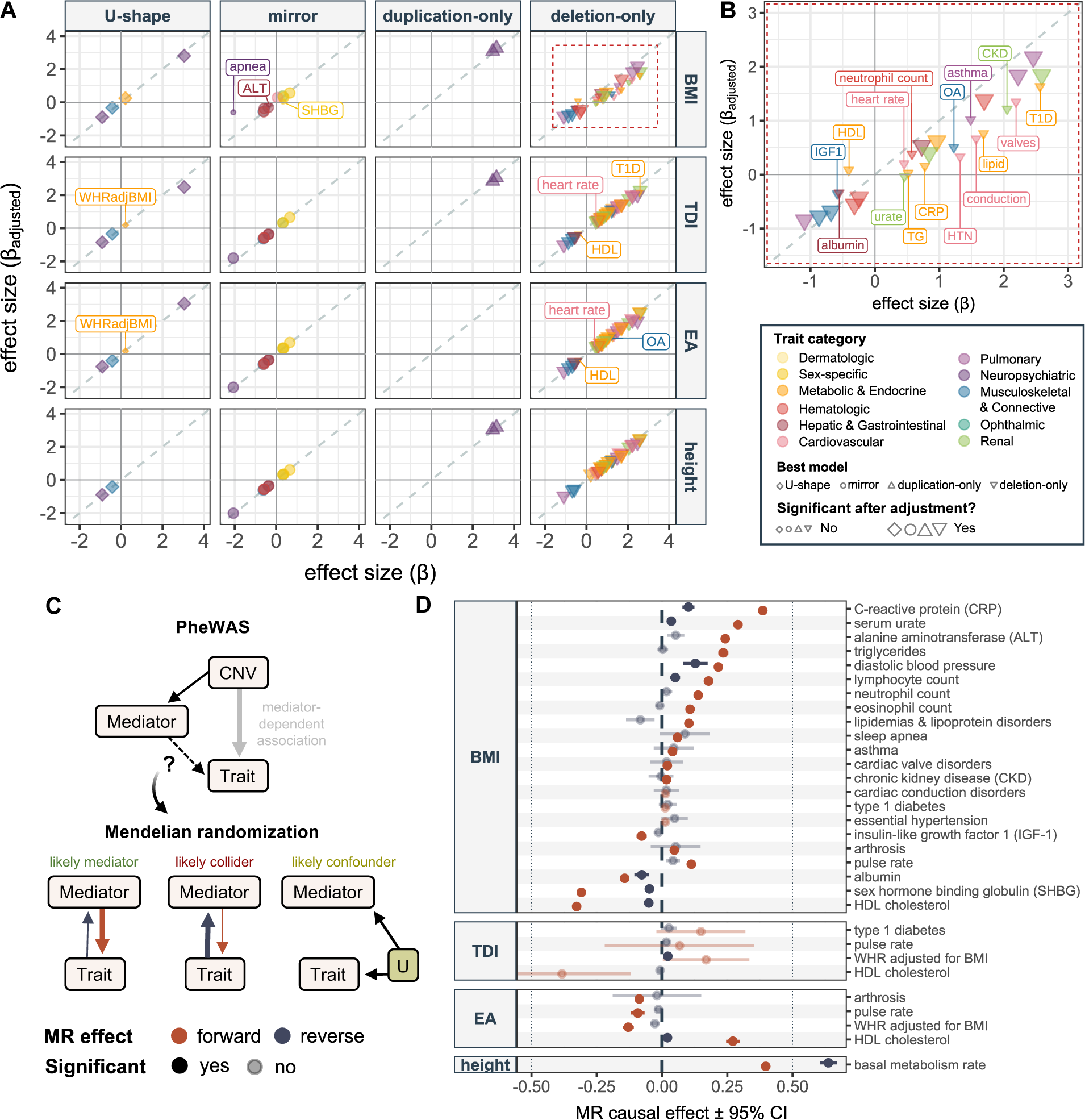
Mediation of 16p11.2 BP4-5 pleiotropy through anthropometric traits and socio-economic factors. (**A**) Effects (beta) of 16p11.2 BP4-5 CNVs on traits with adjustment for potential mediators (y-axis) – i.e., body mass index (BMI), Townsend deprivation index (TDI), age at end of education (EA), and height (rows, right) – against those without adjustment (x-axis), stratified (columns, top) according to the best (i.e., most significant) association model (shape). Only associations that were significant prior to or become significant after adjustment are plotted. Traits are colored according to physiological system. Size reflects whether the effect is Bonferroni significant after adjusting for the potential mediator (large) or not (small). Traits losing significance upon adjustment are labeled. Grey dashed diagonal represents the identity line. (**B**) Enlargement of the area delimited by a red dashed rectangle in (A), showing the effect of BMI adjustment for deletion-driven association, using the same legend as in (A). (**C**) Schematic of the links between copy-number variant (CNV), potential mediators, and assessed traits. Covariate-adjusted phenome-wide association studies (PheWAS) identified CNV-trait associations that are dependent on either of the four tested factors (thick grey arrow) in (A). This scenario can be through mediation, collider bias, or confounding. We used Mendelian randomization (MR) to assess the genetically determined causal effect of the putative mediator on the trait (forward effect, red arrow) and of the trait on the mediator (reverse effect; dark blue arrow). MR effect arrows are proportional to causal effect sizes. When the forward effect is larger than the reverse one, mediation is a likely scenario; when the reverse effect is larger, the putative mediator likely acts as a collider; absence of causal effects likely indicates presence of an unobserved confounder, U. Depending on the scenario, adjustment for the mediator in the regression analysis might (green) or might not (red) be appropriate, as reflected by the color of each scenario’s title. (**D**) Bidirectional forward (red) and reverse (dark blue) MR effects with 95% confidence interval (CI; x-axis truncated on the right) of potential mediators (left y-axis) on traits (right y-axis) for all mediator-trait pairs that either gained or lost significance upon adjustment for the mediator. Non-significant effects (p > 0.05/62 = 8.1 x 10^−4^) are semi-transparent. ALT = alanine aminotransferase; CKD = chronic kidney disease; CRP = C-reactive protein; HDL = high-density lipoprotein cholesterol; HTN = essential hypertension; IGF-1 = insulin-like growth factor 1; OA = arthritis; SHBG = sex hormone binding globulin; T1D = type 1 diabetes; TG = triglycerides; WHRadjBMI = waist-to-hip ratio adjusted for BMI.

Upon adjustment for BMI, TDI, EA, and height, nineteen, four, four, and zero CNV-trait associations fell below the significance cutoff (p ≤ 0.05/117 = 4.3 x 10^−4^), respectively. Comparing effect sizes, only the mirror association with sleep apnea was nominally significantly reduced upon adjustment for BMI (p = 0.04). Remarkably, the association with basal metabolic rate (deletion-only) became significant upon adjustment for height, while the one with diastolic blood pressure (mirror), eosinophil count (deletion-only), and lymphocyte count (deletion-only) became so upon adjustment for BMI (Figure S1A), even though the change in effect size were not significant (p > 0.45). The impact of adjusting for BMI was most striking on deletion-driven associations, for which 61% (16/26) of the associations fell below the significance threshold (Figure 3B). In line with expectations, BMI-dependent traits tended to have a stronger correlation with BMI than those that remained significant upon adjustment for BMI (p = 0.05) (Figure S1B). Among the lost associations, we find nine out of the ten metabolic and cardiovascular traits associated with the deletion. These associations likely reflect secondary consequences of the propensity for obesity of deletion carriers as they include levels of serum lipid and the inflammation biomarker C-reactive protein (CRP), cardiac valve and conduction disorders, and hypertension. The effect of BMI on musculoskeletal, pulmonary, or renal traits is more balanced, with some associations, such as the ones with arthritis (OA), asthma, or urate and chronic kidney disease (CKD), appearing to be driven by BMI, while others, such as grip strength, chronic obstructive pulmonary disease (COPD), or cystatin C and acute kidney injury (AKI), remaining significant upon BMI adjustment. The mediating role of TDI and EA was much milder, as only four associations were lost upon adjustment for either variable – including a shared association with WHR adjusted for BMI, heart rate, and high-density lipoprotein (HDL) cholesterol – suggesting that TDI and EA capture partially overlapping mediatory processes. Surprisingly, associations with psychiatric disorders were not affected by EA, suggesting that cognition and psychiatric diseases are regulated by (at least partially) independent pathways. Finally, the observation that no associations were affected by adjusting for height confirms that the decrease in traits such as grip strength and forced vital capacity among deletion carriers is not driven solely by their short stature.

One caveat of our analysis is that it cannot distinguish whether changes in CNV-trait associations are indeed secondary effects of the mediator on the trait. At least three scenarios could result in the loss (or gain) of a CNV-trait effect upon covariate adjustment (Figure 3C). The first one is mediation, wherein the CNV affects the trait through the mediator, resulting in a dominant causal effect of the mediator on the trait. The second scenario is when the variable we adjusted for turns out to be a collider of the CNV and the trait, in which case we expect a dominant causal effect from the trait to the “mediator”. Finally, data could be explained by an unobserved confounder that affects both the adjustment variable and the trait, in which case we do not expect any causal link between trait and mediator. Of note, in the latter scenario, we further distinguish between whether the CNV has an impact on the confounder, the “mediator”, the trait, or a combination thereof. Importantly, adjusting for the mediator in the regression model is an appropriate solution to obtain meaningful direct CNV-trait effects (i.e., genuine direct pleiotropy) only in the i) mediator scenario or ii) the confounder scenario where the CNV has a direct effect on the trait, in which case adjustment for the mediator could result in a gain of power (Figure 3C). To identify cases where mediation is a likely scenario, we resorted to bidirectional Mendelian randomization (MR), a causal inference approach that allows to estimate the genetically determined causal effect of an exposure on an outcome (Figure 3D; Table S3). Firstly, we estimated the forward mediator-to-trait effect for all 31 mediator-trait pairs that either gained (N = 4; Figure S1A) or lost (N = 27; Figure 3A-B) significance upon adjustment for the mediator. Except for the four TDI-dependent associations which had large confidence intervals due to the lack of good genetic instruments for TDI and the effect of BMI on hypertension, type 1 diabetes, and cardiac conduction disorders, all effects were significant (p ≤ 0.05/62 = 8.1 x 10^−4^), confirming that the mediators can causally influence the involved traits. Secondly, we estimated the reverse trait-to-mediator causal effects. Ten reverse effects were significant and thus represent mediator-trait pairs at risk for collider bias. Yet, for nine of them, the forward effect had a larger magnitude, making the mediator-to-trait causal path more likely. The only exception is the association between the deletion and basal metabolic rate that became significant upon adjustment for height and for which the reverse effect was stronger than the forward effect. This suggests that height could act as a collider and adjustment for it could bias estimates. Hence, we conclude based on the unadjusted effect that the association between the deletion and basal metabolic rate is non-significant. It is also worth noting that six out of seven associations lacking a significant forward effect also lacked a significant reverse effect, possibly indicating presence of an unobserved confounder. This is particularly likely for the BMI effect on hypertension, type 1 diabetes, and cardiac conduction disorders, where estimates are close to null despite being well-instrumented (≥ 50 instruments). Globally, these analyses support that a large fraction (74%) of the flagged associations are likely indirect consequences of the CNV’s effect on our selected mediators.

Next, we focused on the 22 traits whose association with the CNV remained significant after adjusting for BMI, height, TDI, or EA. To confirm that these represent cases of genuine direct pleiotropy, we used a matched-control approach that offers the advantage of allowing adjustment for multiple mediatory variables at once but at the cost of losing some statistical power. Specifically, for each of the 58 deletion and 61 duplication carriers with sufficient data to carry out the matching, we identified individuals with matched age (± 2.5 years), sex (identical), BMI (± 2.5 kg/m^2^), TDI (± 2), income class (identical), and EA (± 1 year) among a pool of copy-neutral, unrelated, white British UKBB participants (Figure S2). For 49 deletion and 60 duplication carriers, at least 25 matched controls could be identified, and phenotype mean or disease prevalence between the two CNV groups and their respective controls were compared (Figure 4; Tables S4-5). Eleven traits (50%) retained a strictly significant effect (p ≤ 0.05/22 = 2.3 x 10^−3^), affecting six independent physiological systems: musculoskeletal, neuropsychiatric, pulmonary/immune, renal, hepatic, and hematological. Specifically, deletion carriers presented with decreased hand grip strength (p = 1.4 x 10^−3^; Figure 4A), shorter stature (p = 1.2 x 10^−5^; Figure 4B), increased alkaline phosphatase (ALP; p = 1.8 x 10^−3^; Figure 4G), decreased forced vital capacity (FVC; p = 2.2 x 10^−3^; Figure 4R), and increased risk for pneumonia (p = 3.8 x 10^−4^; Figure 4Q) and AKI (p = 2.9 x 10^−4^; Figure 4T). Duplication carriers showed decreased bone mineral density (p = 6.3 x 10^−4^; Figure 4C), lower aspartate aminotransferase (AST; p = 1.5 x 10^−3^; Figure 4E) and gamma-glutamyltransferase (GGT; p = 2.2 x 10^−4^; Figure 4F) levels, and reduced fluid intelligence (p = 1.6 x 10^−3^; Figure 4I). Noteworthy is the strong mirror effect on platelet count (Figure 4P), with higher (p = 1.9 x 10^−3^) and lower (p = 3.4 x 10^−4^) counts observed in deletion and duplication carriers, respectively. Whereas for the other phenotypes the other CNV type did not meet the strict significance criteria, all effects, showed a trend for a mirror effect, except for fluid intelligence and AKI, which showed a U-shape trend. Besides reinforcing its long-established consequence on cognitive function, our results assert the role of the hepatic, musculoskeletal, and pulmonary systems in the 16p11.2 BP4-5 pathophysiology through mechanisms that are independent of the CNV’s impact on anthropometric and socio-economic traits.

**Figure 4.**
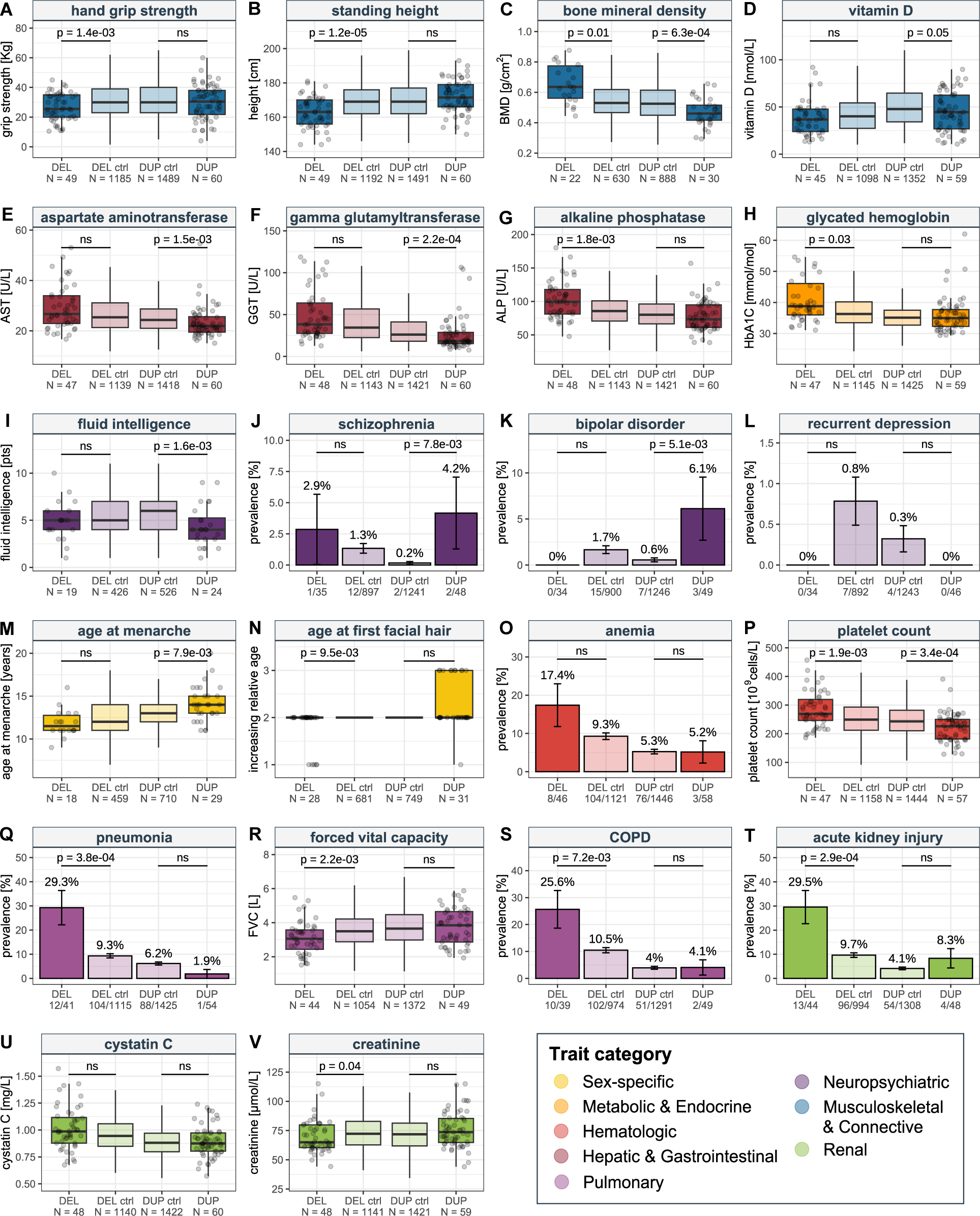
16p11.2 BP4-5 CNV carriers matched-control analyses. (**A**-**V**) Comparison between deletion (DEL) and duplication (DUP) carriers (darker shade) and their respective matched controls (DEL or DUP ctrl; lighter shade) for 22 traits that remained Bonferroni-significant after individually adjusting for body mass index (BMI), height, Townsend deprivation index (TDI), and age at end of education (EA) in Figure 3. For quantitative traits, data are represented as boxplots without outliers and data points for CNV carriers are shown as grey dots. Sample size of each group is indicated as N. P-values of two-sided t-tests comparing CNV carriers to matched controls are indicated. For binary traits, bars represent disease prevalence in percentage and error bars represent the standard error. Number of cases and total sample size for each group is indicated. P-values of two-sided Fisher tests comparing CNV carriers to matched controls are indicated. “ns” indicates p > 0.05. Traits are colored according to physiological systems. COPD = chronic obstructive pulmonary disease.

Finally, we performed sensitivity analyses to validate the robustness of our conclusions. As a negative control, we performed the matched-control analysis for the 24 traits that were significantly associated with 16p11.2 BP4-5 CNVs in our PheWAS but whose association was dependent on adjustment for mediators or that could not be tested in the covariate analysis due to high trait-mediator correlation (Figure S3; Tables S4-5). In line with these associations being secondary consequences to the effect of the CNV on factors on which the matching was performed, only three traits had a nominally significant CNV association, and none survived Bonferroni correction. This strongly contrasts with our main matched-control analysis, where only three traits *lacked* a nominally significant effect: recurrent depression (Figure 4L), anemia (Figure 4O), and cystatin C (Figure 4U). This absence of results could either be the result of a loss in statistical power resulting from CNV carrier subsampling or by these associations being driven by a combination of factors on which the matching was performed. The former could be exacerbated by the fact that CNV carriers with the more extreme phenotypes were less likely to have 25 matched controls in the UKBB. To explore this hypothesis, we compared mean trait value or disease prevalence between the subset of CNV carriers used for the matched-control analysis and the one excluded due to missing data or lack of a sufficient number of matched controls (Figure S4; Tables S4-5). Except for recurrent depression and FVC, all comparisons were non-significant (p ≥ 0.05), indicating that subsampling does not strongly impact our results. For recurrent depression, the only three duplication carriers diagnosed with the disease were not included in the matched-control analysis (p = 0.03; Figure S4L), indicating that the non-significant effect of the duplication on recurrent depression (Figure 4L) is likely caused by subsampling. For FVC, excluded deletion carriers exhibited a more pronounced phenotypic decrease than the ones retained for the matched-control analysis (p = 0.02; Figure S4R), suggesting that an even more extreme difference would have been observed if these individuals had been included in the matched-control analysis (Figure 4R). Conversely, the role of the CNV on anemia risk and cystatin C is likely driven by the effect of the CNV on adiposity and socio-economic status.

## Discussion

In this study, we perform a comprehensive PheWAS assessing the relation between 16p11.2 BP4-5 CNVs and 117 complex traits and diseases in the general population through four dosage mechanisms of action. Our results confirm the extreme pleiotropy of 16p11.2 BP4-5 rearrangements, with 46 traits associating with the CNV. In line with the more deleterious nature of the deletion, haploinsufficiency associated with 38 unique traits, while only 10 traits associated with the region’s duplication. Further emphasizing how the same genetic region can affect different traits through different dosage mechanisms, we identify traits for which the loss and gain of a copy had an opposite (e.g., BMI or platelet count) or alternatively, a similar (e.g., grip strength or fluid intelligence) consequences on the phenotype. Besides assessing the role of dosage in pleiotropy, we also estimated the fraction of associations that are likely to be secondary to some hallmark features of the CNV and validated through bidirectional MR that mediation is a likely scenario. While height did not mediate any associations, sixteen (61%) of the deletion-driven associations were found to be BMI-dependent, thirteen of which (81%) received support from MR for a scenario wherein the association is consequential to an initial increase in BMI. Conversely, the role of EA and TDI was more subtle, with only five associations showing confounding by these factors. Importantly, some associations were found to be independent of all the tested mediators, suggesting genuine direct pleiotropy of the region on musculoskeletal, hepatic, metabolic, neuropsychiatric, reproductive, hematological, pulmonary, immune, and renal function.

Our findings have far-reaching consequences for clinical practice and highlight knowledge gaps. First, our results show that increased BMI in deletion carriers drives numerous adult-onset comorbidities. Studies have shown that weight gain in 16p11.2 BP4-5 deletion carriers starts during early childhood, to rapidly progress to obesity^9,16,45–47^. This emphasizes the importance of following pediatric cases by a dedicated team of endocrinologists and nutritionists who can implement a weight control strategy at an early age to attenuate ensuing adult co-morbidities. Second, we show that some other traits are affected independently of the CNV’s effect on BMI, cognition, and SES. Besides recapitulating well-established hallmark features, such as the CNV’s negative impact on cognitive ability or the duplication-specific risk of bipolar disorder or depression, we also link the CNV with milder afflictions of systems that had previously been implicated in clinical cohorts. For instance, increased risk for AKI might be the consequence of subclinical structural defects of the kidney that could affect renal function in the long term, paralleling the predisposition of deletion carriers to congenital anomalies of the kidney and urinary tract^48–50^. Similarly, increased risk for pneumonia might reflect an impaired immune system that is exacerbated into a full-blown immunodeficiency in deletion carriers that also present with a loss-of-function variant in *CORO1A*^51^ (MIM: 605000). Other traits that are affected through BMI-independent mechanisms, such as bone mineral density, platelet counts, pulmonary function, and liver enzymes have not been linked with the CNV in clinical cohorts and future research should establish how often these traits are altered in carriers and which are the molecular mechanisms that mediate this pleiotropy. These could be explored by gene-to-trait mapping strategies such as rare variant gene burden tests^52,53^, as well as MR^54^ or colocalization^55^ that integrate association signals from common SNP-GWAS with transcriptomic and proteomic data to pinpoint genes linked with specific phenotypes. These data could also be leveraged to generate gene-by-trait association matrices whose clustering may reveal groups of traits with shared underlying genetic influences and for which CNV associations are more likely to disappear upon adjustment for one another. Thirdly, our results expose intriguing findings, casting light on questions that remain unanswered by the current study. For instance, the BMI-dependent association of the deletion with type 1 diabetes could be driven by misdiagnosing type 2 diabetes as type 1 due to early-onset diabetes following early-onset obesity. We also identify an association between the deletion and decreased creatinine levels. Creatinine levels are typically *elevated* in patients with renal dysfunction, as is the case for many deletion carriers. We speculate that these results could be the consequences of reduced hepatic function or muscle mass, both of which are present among deletion carriers. Similarly, it remains unclear whether elevated levels of ALP – for which levels of specific isoenzymes were not determined in UKBB – reflect hepatic, renal, or skeletal dysfunction. Validation of these hypotheses requires in-depth phenotypic characterization of carriers’ medical records but will be crucial to better define the molecular pathophysiology of 16p11.2 BP4-5 CNV carriers and hopefully lead to actionable insights related to the management of the condition’s co-morbidities.

Our study is not without limitations. First, by assessing a relatively homogenous cohort, our study likely misses pleiotropic consequences that are only expressed in certain genetic or environmental backgrounds, a phenomenon exacerbated by the relatively small absolute number of CNV carriers which hinders our statistical power. Future studies are needed to confirm trends that we observe at sub-significant level. Second, we decided to focus on only four covariates, which based on the literature, represent strong candidates to mediate indirect pleiotropic consequences of the region’s rearrangement. While height and BMI can be measured with relatively high accuracy, EA and TDI only offer rough and imperfect proxies for complex characteristics such as cognitive function and SES, possibly explaining their weaker mediatory role. Other factors that we did not assess might mediate the relation between 16p11.2 BP4-5 CNVs and some of the associated traits. Third, the conducted MR analysis comes with its own limitations, namely violation of the exclusion-restriction assumption via correlated pleiotropy, which may have resulted in false positive mediator-to-trait causal effects^56,57^. Still, if both adjusted and unadjusted regression analyses show a significant CNV effect, we can convincingly suggest that independent pleiotropic mechanisms are at play. Finally, while our study brings us a step closer to understanding the pleiotropy of the region, it fails to provide molecular insights into mechanisms of pleiotropy, for which experimental approaches and leveraging of other mutational classes offer promising avenues.

In conclusion, our study provides a framework to start disentangling the complex pleiotropic patterns associated with genomic disorders. For 16p11.2 BP4-5, the latter appears to be a mixture of indirect effects mediated by the impact of the CNV on adiposity and cognition, and direct effects on a broad range of physiological systems. This suggests that independent molecular mechanisms are involved in translating dosage changes into the many co-morbidities linked to the genomic disorder.

## Data Availability

UK Biobank data are available for registered users and were accessed through the application number 16389. Summary statistics used for Mendelian randomization studies are publicly available, as described in the Methods. All data produced by this study are available in Supplemental Tables. Code will be made available upon publication at: https://github.com/cauwerx/16p11.2_BP4-5_pleiotropy.

## Abbreviations

ALP: alkaline phosphatase
ALT: alanine aminotransferase
AKI: acute kidney injury
AST: aspartate aminotransferase
BMI: body mass index
BP: breakpoint
CI: confidence interval
CKD: chronic kidney disease
CNV: copy-number variants
COPD: chronic obstructive pulmonary disorder
CRP: C-reactive protein
EA: educational attainment
FVC: forced vital capacity
GGT: gamma-glutamyltransferase
GWAS: genome-wide association studies
HbA1c: glycated hemoglobin
HDL: high-density lipoprotein
HTN: essential hypertension
ICD-10: International Classification of Diseases, 10^th^ Revision
IGF-1: insulin-like growth factor 1
kb: kilobase pair
MR: Mendelian randomization
OA: arthritis
PheWAS: phenome-wide association study
SE: standard error
SES: socioeconomic status
SHBG: sex hormone binding globulin
SNP: single nucleotide polymorphism
T1D: type 1 diabetes
TDI: Townsend deprivation index
UKBB: UK Biobank
WHR: waist-to-hip ratio
WHRadjBMI: WHR adjusted for BMI.

## Author contributions

CA performed all analyses, except for MR analyses conducted by SM; ZK supervised statistical analyses; CA, ZK, and AR interpreted the data; CA generated the figures and drafted the manuscript; ZK and AR made critical revisions; All authors approved the final manuscript.

## Acknowledgments

We thank UKBB biobank participants for sharing their data. Computations were performed on the Urblauna server from the University of Lausanne. The study was funded by the Swiss National Science Foundation (31003A_182632, AR; 310030_189147, ZK), Horizon2020 Twinning projects (ePerMed 692145, AR), and the Department of Computational Biology (ZK) and the Center for Integrative Genomics (AR) from the University of Lausanne.

