## Supplemental Figures for "Disentangling mechanisms behind the pleiotropic effects of proximal 16p11.2 BP4-5 CNVs"

**Figure S1.** Adjustment for potential mediators of 16p11.2 BP4-5 pleiotropy.

**Figure S2.** Number of matched controls per 16p11.2 BP4-5 CNV carrier.

**Figure S3.** 16p11.2 BP4-5 CNV carriers matched-control analyses negative controls.

**Figure S4.** Impact of CNV carriers subsampling on matched-control analyses.

**SUPPLEMENTAL FIGURES**

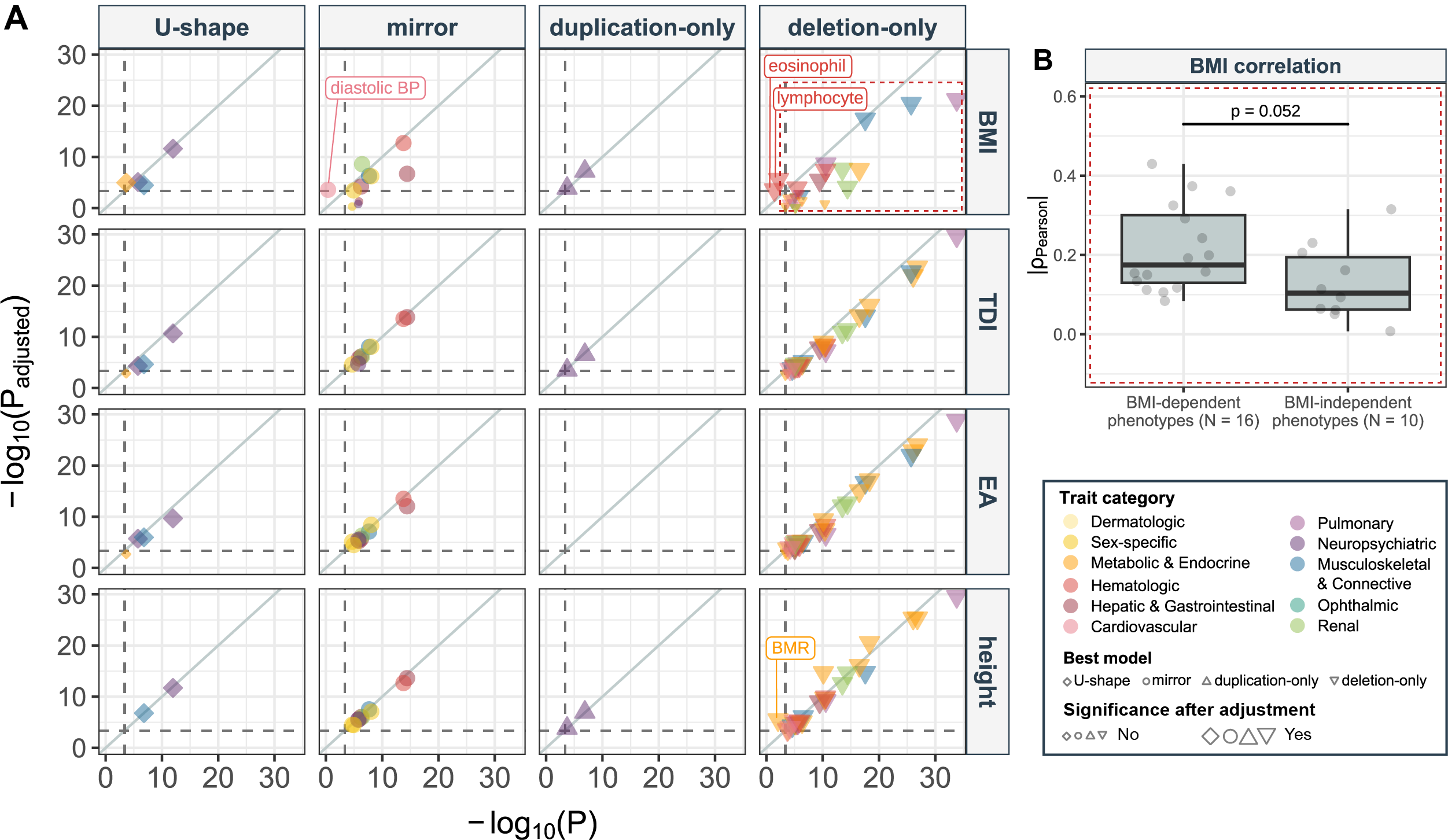

**Figure S1. Adjustment for potential mediators of 16p11.2 BP4-5 pleiotropy.**

(**A**) Negative logarithm of p-values of 16p11.2 BP4-5 CNV effect on traits with adjustment for potential mediators (y-axis) – i.e., body mass index (BMI), Townsend deprivation index (TDI), age at end of education (EA), and height (rows, right) – against those without adjustment (x-axis), stratified (columns, top) according to the best (i.e., most significant) association model (shape). Only associations that were significant prior to or become significant after adjustment are plotted. Traits are colored according to physiological systems. Size reflects whether the effect is Bonferroni significant after adjusting for the potential mediator (large) or not (small). Traits that become Bonferroni significant after adjustment are labeled. Grey diagonal represents the identity line; Dark grey dashed lines represent the Bonferroni threshold of p ≤ 0.05/117 = 4.3 x 10^-4^. (**B**) Pearson correlation of BMI with traits that are significantly associated with the deletion (red dashed square in (A)), stratified according to whether the association with the deletion is lost (“BMI-dependent) or not (“BMI-independent”) after adjustment for BMI. The P-value compares the two groups with a two-sided t-test. Number of traits in the two groups is indicated as N. BMR = basal metabolic rate; BP = blood pressure; eosinophil = eosinophil count; lymphocyte = lymphocyte count.

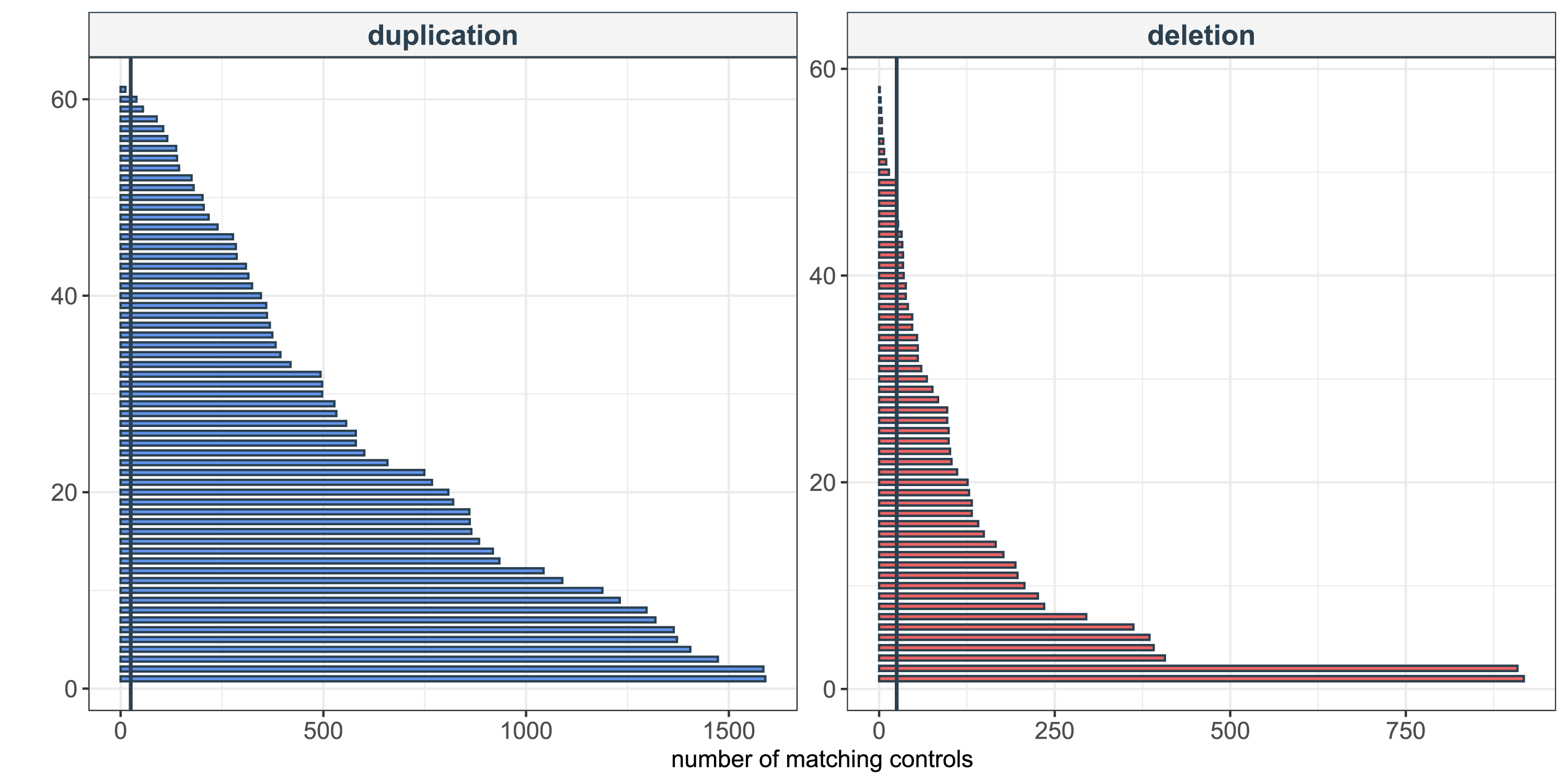

**Figure S2. Number of matched controls per 16p11.2 BP4-5 CNV carrier.**

Total number of identified matched controls (x-axis) per 16p11.2 BP4-5 duplication (N = 61; blue; left) and deletion (N = 58; red; right) carrier (y-axis). The black vertical line represents the cutoff of 25 randomly sampled matched controls per CNV carrier. In total 60 duplication and 49 deletion carriers passed this threshold.

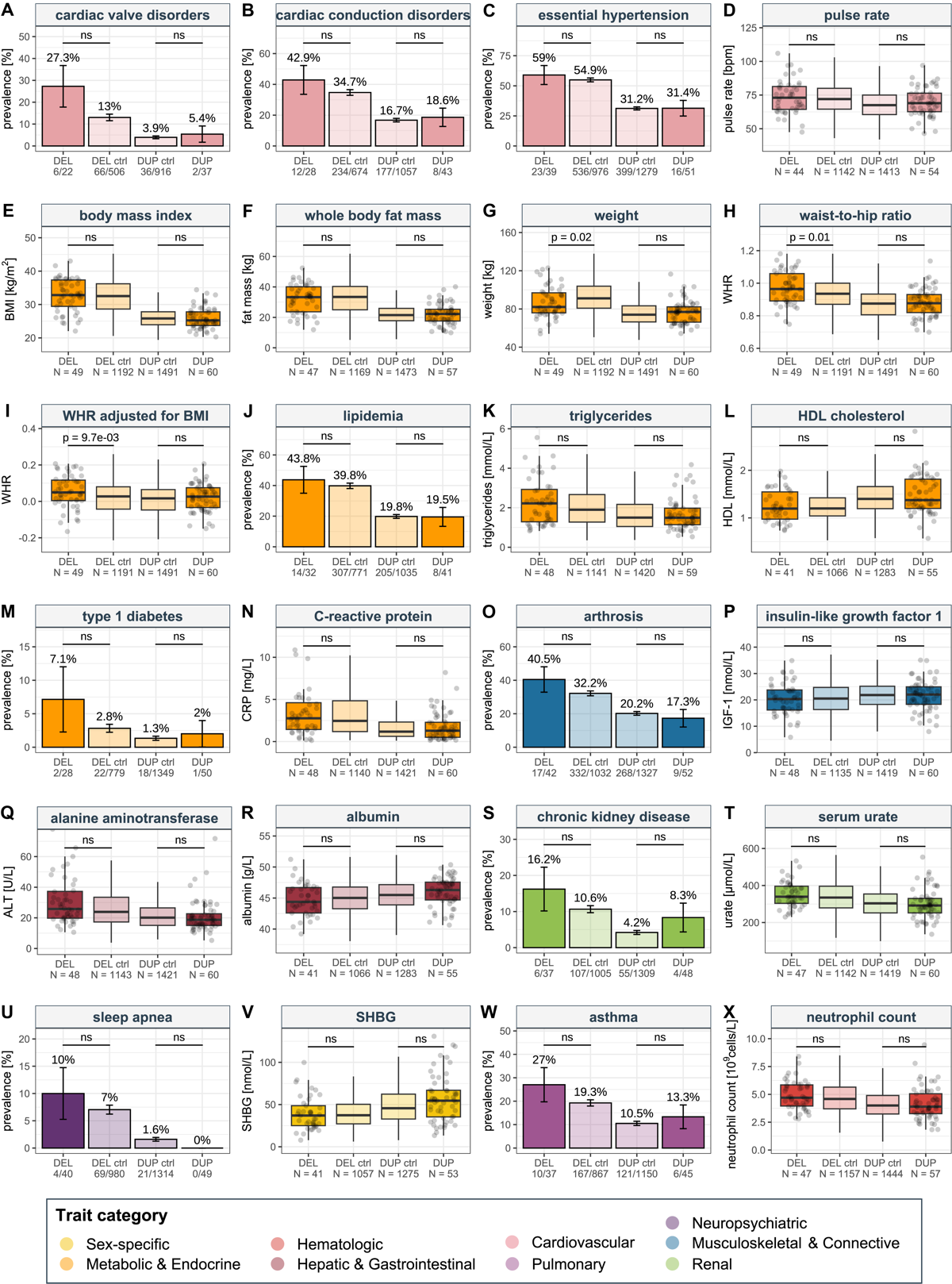

**Figure S3. 16p11.2 BP4-5 CNV carriers matched-control analyses negative controls.**

(**A**-**X**) Comparison between deletion (DEL) and duplication (DUP) carriers (dark shade) and their respective matched controls (DEL or DUP ctrl; lighter shade) for 24 traits that were significantly associated with 16p11.2 BP4-5 CNVs in our PheWAS but whose association was dependent on adjustment for mediators or that could not be tested in the covariate analysis due to high trait-mediator correlation. For quantitative traits, data are represented as boxplots without outliers and data points for CNV carriers are shown as grey dots. Sample size of each group is indicated as N. P-values of a two-sided t-test comparing CNV carriers to matched controls are indicated. For binary traits, bars represent disease prevalence in percentage and error bars represent the standard error. Number of cases and total sample size for each group are indicated. P-values of two-sided Fisher tests comparing CNV carriers to matched controls are indicated. “ns” indicates p > 0.05. Traits are colored according to physiological systems. SHBG = sex hormone binding globulin.

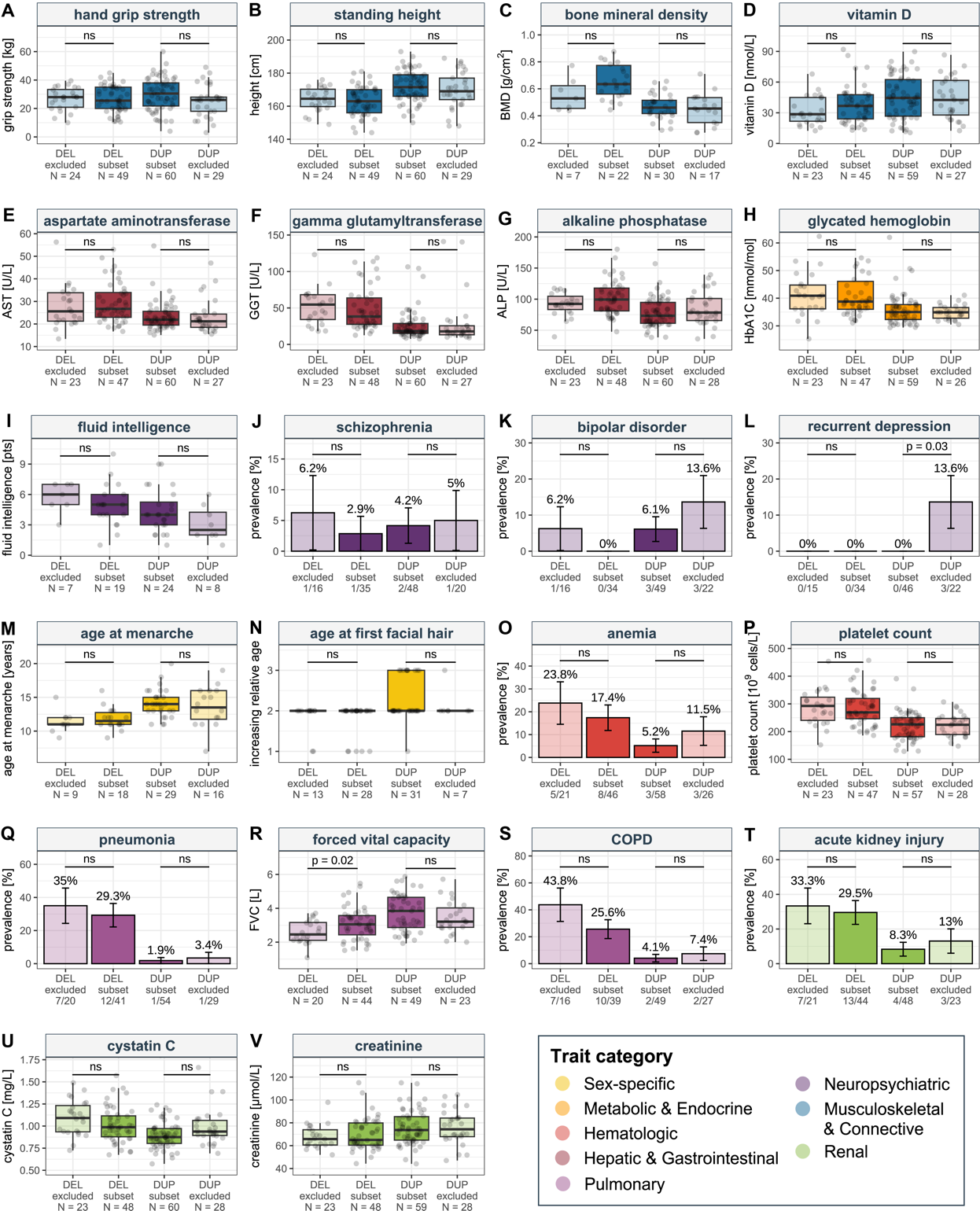

**Figure S4. Impact of CNV carriers subsampling on matched-control analyses.**

(**A**-**V**) Comparison between deletion (DEL; N_max_ = 49) or duplication (DUP; N_max_ = 60) carriers that were in the subset (subset; darker shade) used for the matched-control analyses against deletion (DEL; N_max_ = 24) or duplication (DUP; N_max_ = 29) carriers that were not included due to lack of data (excluded; lighter shade) for 22 traits which remained Bonferroni-significant after adjusting for body mass index (BMI), height, Townsend deprivation index (TDI), and age at end of education (EA) in Figure 3. For quantitative traits, data are represented as boxplots without outliers and data points for CNV carriers are shown as grey dots. Sample size of each group is indicated as N. P-values of two-sided t-test comparing CNV carriers to matched controls are indicated. For binary traits, bars represent disease prevalence in percentage and error bars represent the standard error. Number of cases and total sample size for each group is indicated. P-values of two-sided Fisher tests comparing CNV carriers to matched controls are indicated. “ns” indicates p > 0.05. Traits are colored according to physiological systems. COPD = chronic obstructive pulmonary disease.
